# Clinical and Virological Features of patients hospitalized with different types of COVID-19 vaccination in Mexico City

**DOI:** 10.1101/2022.05.06.22274772

**Authors:** Alejandra Hernández-Terán, Pamela GarciaDiego, Marco Villanueva-Reza, Celia Boukadida, Blanca Taboada, Eduardo Porras, Victor Ahumada-Topete, Kathia Tapia Diaz, Margarita Matias-Florentino, Marissa Pérez-García, Santiago Ávila-Rios, Fidencio Mejía-Nepomuceno, Ricardo Serna-Muñoz, Fortunato Juárez-Hernández, Maria Eugenia Jiménez-Corona, Eduardo Becerril-Vargas, Omar Barreto, Martínez-Orozco Jose Arturo, Rogelio Pérez-Padilla, Carlos F. Arias, Joel Armando Vázquez-Pérez

## Abstract

Coronavirus disease 2019 (COVID-19) vaccines are very effective at protecting against severe disease and death. However, the impact of the vaccine used, viral variants, and host factors on disease severity remain poorly understood. Here we compared COVID-19 clinical presentations and outcomes in vaccinated and unvaccinated patients in Mexico City. From March to September 2021, clinical and demographic characteristics were obtained from 1,014 individuals with a documented SARS-CoV-2 infection, and viral variants were identified in a subset of 386 patients. We compared unvaccinated, partially vaccinated, and fully vaccinated patients, stratifying by age groups. We fitted multivariate statistical models to evaluate the impact of vaccination status, SARS-CoV-2 lineages, vaccine types, and clinical parameters. Most hospitalized patients were unvaccinated. In patients over 61 years old, mortality was significantly higher in unvaccinated compared to fully vaccinated individuals. In patients aged 31 to 60 years, vaccinated patients were more likely to be outpatients (46%) than unvaccinated individuals (6.1%). We found immune disease and age above 61 years old as risk factors. While fully vaccination was found as the most protective factor against in-hospital death. This study suggests that vaccination is essential to reduce mortality in a comorbid population such as that of Mexico.

## INTRODUCTION

Severe acute respiratory syndrome coronavirus 2 (SARS-CoV-2) was first detected in Wuhan, China, in December 2019 [1]; since then, more than 500 million people have been infected, and over six million have died worldwide [2]. SARS-CoV-2 has evolved and several mutations throughout the genome have been detected worldwide [3]. Genomic mutations are anticipated events during virus replication, and although most mutations are expected to be neutral some can confer a fitness advantage and be fixed in the viral genome [4,5]. The accumulation of mutations over time generates new SARS-CoV-2 variants [6]. The World Health Organization (WHO) has classified them as variants of concern (VOC), variants of interest (VOI), and variants under monitoring (VUM) (WHO, tracking SARS-CoV-2 variants). VOC are SARS-CoV-2 variants with increased transmissibility, virulence, or decreased effectiveness of vaccines, therapeutics, diagnostics, or public health and social measures [6].

As in other countries, in Mexico, several variants classified as VOC, VOI, and VUM have been detected, particularly Delta, Alpha, Gamma, Mu [7], and more recently Omicron [8]. Moreover, B.1.1.519 was detected as the predominant lineage in Mexico during late 2020 and the first months of 2021 [9]. The B.1.1.519 lineage was dominant during the second COVID-19 wave in Mexico and may have been associated with an increased risk of severe and fatal outcomes [10].

In Mexico, vaccination against COVID-19 began in December 2020 with the health personnel and people older than 60 years old, followed by people over 50. Until September 30, 2021, 101,190,484 doses have been applied with a variety of platforms; of which 43,431,200 doses were of Oxford-AstraZeneca (ChAdOx1 nCov-19 adenoviral vector), 33,022,275 of Pfizer BioNTech (RNA platform, mRNA-BNT162b2), 20,000,000 of SinoVac (inactivated virus), 9,900,000 of Sputnik V (Gam-COVID-Vac), 8,149,930 of CanSino (Convidecia or AD5-nCOV); 3,500,000 of Moderna (mRNA-1273), and 1,350,000 of Janssen (AD26.coV2.s). By the end of September 2021, almost 50% of the population over 18 years old had received at least one dose of vaccine [11].

Importantly, COVID-19 vaccines have been shown to be effective at preventing severe disease and death even in patients with comorbidities [12]. As a result, fully vaccinated persons are less likely to experience severe disease or death than unvaccinated persons with the same medical conditions [13]. Nonetheless, vaccines are less effective at protecting from SARS-CoV-2 infections, and this partial protection seems to wane after a few months, leading to infections in vaccinated individuals (breakthrough infections) [14]. Furthermore, one of the most critical factors behind the changing vaccine effectiveness is the viral variability and the mutations that affect the recognition of neutralizing antibodies elicited by vaccination or previous infections [15,16]. These mutations are located mainly in the Receptor Binding Domain (RBD) of Spike protein [17].

Understanding the impact of vaccination, viral variants, and host factors on disease severity is critical to guide COVID-19 vaccination campaigns and protective measures. Here, we analyzed the main risk factors for severe COVID-19, comparing the outcomes and clinical presentation of vaccinated and unvaccinated patients, and analyzing the effects of both SARS-CoV-2 lineages and vaccine types in a tertiary hospital in Mexico City from March to September 2021.

## MATERIAL AND METHODS

### Study participants and clinical data

We analyzed 1014 patients with SARS-CoV-2 infection who received medical attention from March to September 2021 at the Instituto Nacional de Enfermedades Respiratorias (INER), belonging to the Ministry of Health of Mexico. INER is a reference hospital for respiratory diseases that primarily provide services for uninsured individuals and was designated as a hospital exclusive for COVID-19 in March 2020, offering between 150-200 beds for the attention of critically ill patients.

For all patients, demographic data, clinical symptoms, laboratory, radiology data, and outcome-related information were obtained from electronic medical records. Clinical management was performed according to the standards of care and attending physicians. Follow-up went from the time of admission for up to 28 days. This study was reviewed and approved by the Science, Biosecurity, and Bioethics Committee of the Instituto Nacional de Enfermedades Respiratorias (B-10-20). In addition, the Institution requested informed consent for the recovery, storage, and use of biological remnants for research purposes.

### SARS-CoV-2 diagnostics

Oropharyngeal and/or nasopharyngeal swabs were collected, and the diagnosis was made using validated RT-qPCR protocols for SARS-CoV-2 RNA detection, approved by the World Health Organization (WHO).

### RNA extraction and sequencing

#### Complete genome sequencing

Viral nucleic acid extraction was performed using a MagNa Pure L.C. 2-0 system (Roche, Indianapolis, OH, USA) or QIAamp viral RNA Minikit (Qiagen, Hilden Germany). Libraries for whole-genome sequencing of SARS-CoV-2 were generated using the protocol developed by the ARTIC Network (https://artic.network/2-protocols.html) or a long-amplicon-based method [18]. Libraries were sequenced on a MiSeq sequencing platform using a 2×150-cycle or a NextSeq 500 platform using 2×150-cycle mid-output kits to obtain paired-end reads (Illumina, San Diego, CA, USA). The DRAGEN COVIDSeq Test Pipeline on BaseSpace Sequence Hub performed the analysis, mapping, and consensus.

#### Spike partial sequencing

Partial sequencing of the Spike segment was performed by Sanger sequencing in samples with incomplete genome or samples with cycle threshold (Ct) values above 28. Briefly, 956 bp amplicons (944-1900 nucleotide sequence, 315-633 aa) were obtained using specific primers for SARS-CoV-2: SF3 CTTCTAACTTTAGAGTCCAACC and SR4 GCCAAGTAGGAGTAAGTTGAT. The amplicons were sequenced in both directions with the same primers. Sequencing reactions were performed with BigDye Terminator v3.1 (Life Technologies, Carlsbad, CA) as instructed by the manufacturer. Sequences were obtained by capillary electrophoresis using an ABI Prism 3500 Genetic Analyzer (Life Technologies) and were assembled using MEGA 10.0 [19]. The sequences mentioned above can be found at GenBank (accession numbers: ON158371-ON158443).

### Phylogenetic analysis

To perform a phylogenetic analysis, we analyzed only samples with complete genomes of hospitalized and ambulatory patients from our cohort (N= 311). In addition, for the genomic surveillance, 219 complete Mexican genomes available in the GISAID platform, including 140 sequences of INER from May 2020 to November 2020, 42 and 37 sequences of different States of Mexico from March 2020 to November 2020, and March 2021 to September 2021 respectively were used. Also, we included 10 USA sequences from January 2020 to November 2021 and one Wuhan 2019 sequence for reference. To construct the analysis, we selected only the Spike sequences of these 541 sequences. Sequence alignments were created with MAFFT V7 [20] and edited with MEGA 10.0. A maximum-likelihood tree was constructed for the whole Spike sequence using MEGA 10.0. The General Time-Reversible (GTR) model was selected with 5-parameter gamma-distributed rates and 1,000 bootstrap replicates. Edition of the trees was made using FigTree [21].

### Statistical analyses

To evaluate the impact on clinical outcomes, we divided our cohort into three groups: 1) unvaccinated: defined as those patients who had never received a SARS-CoV-2 vaccine; 2) partially vaccinated: defined as those patients who had not yet completed the standard vaccination schedule (depending on the vaccine) or had received the last dose less than 14 days before symptom onset; and 3) fully vaccinated: defined for those patients who had already completed the vaccination schedule and with more than 14 days after the last dose. As advanced age is a significant driver of severe COVID-19 [22]; we categorized patients into three groups: <30 years old, 31-60 years old, and > 61 years old. In addition, the severity of the disease was categorized according to the NIH COVID-19 treatment guidelines [23] into four groups: 1) Mild illness: individuals who have any of the various signs and symptoms of COVID-19 (e.g., fever, cough, sore throat, malaise, headache, muscle pain, nausea, vomiting, diarrhea, loss of taste, and smell) but who do not have shortness of breath, dyspnea, or abnormal chest imaging, 2) Moderate illness: individuals who show evidence of lower respiratory disease during clinical assessment or imaging and who have and oxygen saturation (SpO_2_) >94% on room air at sea level, 3) Severe illness: individuals who have SpO_2_ <94% on room air at sea level, a ratio of arterial partial pressure of oxygen to fraction of inspired oxygen (PaO_2_/FiO_2_) <300 mmHg, a respiratory rate >30 breaths/min, or lung infiltrates in >50% of the lung fields, and 4) Critical illness: individuals who have respiratory failure, septic shock, or multiple organ dysfunction. All statistical analyses were performed using R v4.0.2 in RStudio v1.3.1 [24] and the packages ggplot2 (v3.3.3) [25], stats (v3.6.2) [24], survival (v3.2.13) [26], and survminer (v0.4.9) [27]. We used stacked bar plots, density plots, pie charts, or heatmaps constructed in the ggplot2 package to represent data proportion. For numerical data representation, we used boxplots constructed in the ggplot2 package. For all cases, categorical variables were statistically compared using Chi-square or Fisher’s exact test and continuous variables using Wilcoxon rank-sum test in R base.

To predict the clinical outcome (deceased vs. recovered) we constructed Generalized Linear Models (GLM) using a Binomial family with a *logit* link in R base. We fitted one model as a function of vaccination status and another model as a function of clinical parameters. Both models were adjusted by comorbidities, age, and sex. Odds ratios and 95% CI for all selected variables were calculated and plotted in the final model. Finally, to detect variables that significantly affect survival probability, we calculated Kaplan-Meier curves coupled with the Cox proportional hazard models to determine factors affecting survival, using the survival package and plotted with the survminer package. We used the hospitalization length in days as a time variable for all curves, the outcome (deceased or recovered) standardized at 28 days as a dependent variable, and the tested variables as exposure. Finally, for both the GLM and the Kaplan-Meier curves, we categorized the numerical variables (e.g., age, D dimer.) into three groups according to the data distribution: 1) all values under the 25th percentile, 2) values between the 25th and 75th percentiles, and 3) values above the 75th percentile.

## RESULTS

### Clinical and demographic characteristics of the cohort

A total of 1014 patients admitted to INER from March to September 2021 positive for SARS-CoV-2 infection were included. 124 (12%) were outpatients, and 890 (88%) were hospitalized due to respiratory failure. 48 patients in the cohort had incomplete information; thus, only 111 out of 124 outpatients and 855 out of 890 hospitalized patients had their clinical and demographic data analyzed (Table 1).

**Table 1.** Demographics of the cohort.

|  | Outpatients (N=111) |  |  |  | Hospitalized (N=855) |  |  |  |
| --- | --- | --- | --- | --- | --- | --- | --- | --- |
|  | Unvaccinated<br>(N=39) [35.1%] | Partially vaccinated<br>(N=32) [28.8%] | Fully vaccinated<br>(N=40) [36%] | P | Unvaccinated<br>(N=572) [66.9%] | Partially vaccinated<br>(N=178) [20.8%] | Fully vaccinated<br>(N=105) [12.2%] | P |
| **Age** |  |  |  |  |  |  |  |  |
| <30, n (%) | 13 (33.3%) | 4 (12.5%) | 14 (35%) | 0.03 | 54 (9.4%) | 1 (0.5%) | 0 | <0.001 |
| 31-60, n (%) | 25 (64.1%) | 22 (68.7%) | 18 (45%) | 0.04 | 382 (66.7%) | 132 (74.1%) | 21 (20%) | <0.001 |
| =/≥60, n (%) | 1 (2.50%) | 6 (18.7%) | 8 (20%) | 0.03 | 135 (23.6%) | 45 (25.2%) | 82 (78%) | <0.001 |
| NA | 0 | 0 | 0 |  | 1 | 0 | 2 |  |
| **Gender** |  |  |  |  |  |  |  |  |
| Female, n (%) | 17 (43.5%) | 20 (62.5%) | 16 (40%) | ns | 200 (34.9%) | 65 (36.5%) | 52 (49.5%) | 0.02 |
| NA | 0 | 0 | 0 |  | 0 | 0 | 0 |  |
| **Vaccine type** |  |  |  |  |  |  |  |  |
| Total of vaccinated patients |  | 32 (100%) | 40 (100%) |  |  | 179 (100%) | 104 (100%) |  |
| Pfizer, n (%) | na | 12 (37%) | 28 (70%) | 0.01 | na | 34 (19%) | 14 (13.3%) | ns |
| AztraZeneca, n (%) | na | 13 (40.6%) | 4 (10%) | 0.02 | na | 69 (38.7%) | 15 (14.2%) | <0.001 |
| SinoVac, n (%) | na | 3 (9.3%) | 5 (12%) | ns | na | 18 (10.1%) | 35 (33.3%) | <0.001 |
| Sputnik, n (%) | na | 2 (6.2%) | 2 (5%) | ns | na | 51 (28.6%) | 21 (20%) | ns |
| Cansino, n (%) | na | 0 | 1 (2.5%) | na | na | na | 18 (17.1%) | na |
| J&J, n (%) | na | 0 | 0 | na | na | 1 (0.56%) | 0 | ns |
| Moderna, n (%) | na | 0 | 0 | na | na | 0 | 1 (0.95%) | ns |
| NA | na | 2 | 0 |  | na | 5 | 1 |  |
| **Comorbidities** |  |  |  |  |  |  |  |  |
| Diabetes, n (%) | 2 (5.1%) | 4 (12.5%) | 1 (2.5%) | ns | 115 (20.1%) | 44 (24.7%) | 42 (40%) | <0.001 |
| Hypertension, n (%) | 5 (12.8%) | 4 (12.5%) | 5 (12.5%) | ns | 147 (25.6%) | 45 (25.2%) | 53 (53.3%) | <0.001 |
| Obesity, n (%) | 5 (12.8%) | 7 (21.8%) | 4 (10%) | ns | 255 (44.5%) | 82 (46%) | 47 (44.7%) | ns |
| Smoking, n (%) | 2 (5.1%) | 1 (3.1%) | 2 (5%) | ns | 177 (30.9%) | 57 (32%) | 25 (23.8%) | ns |
| COPD, n (%) | 0 | 0 | 2 (5%) | na | 12 (2%) | 2 (1.1%) | 4 (3.8%) | ns |
| Immune disease, n (%) | 0 | 0 | 2 (5%) | na | 12 (2%) | 1 (0.5%) | 3 (2.8%) | ns |
| NA | 89 | 85 | 105 |  | 33 | 13 | 10 |  |
| **Number of comorbidities** |  |  |  |  |  |  |  |  |
| None, n (%) | 22 (56.4%) | 13 (40.6%) | 25 (62.1%) | 0.05 | 97 (16.9%) | 34 (19.1%) | 10 (9.5%) | 0.05 |
| 1, n (%) | 9 (23.07%) | 2 (6.2%) | 4 (10%) | ns | 217 (37.9%) | 58 (32.5%) | 21 (20%) | <0.001 |
| 2, n (%) | 2 (5.1%) | 1 (3.1%) | 4 (10%) | ns | 170 (29.7%) | 55 (30.8%) | 35 (33.3%) | ns |
| =/≥ 3, n (%) | 1 (2.5%) | 3 (9.3%) | 2 (5%) | ns | 87 (15.2%) | 31 (17.4%) | 39 (37.1%) | <0.001 |
| NA | 5 | 13 | 5 |  | 1 | 0 | 0 |  |
| **In-hospital treatment** |  |  |  |  |  |  |  |  |
| Dexamethasone, n (%) | 5 (12.8%) | 2 (6.2%) | 1 (2.5%) | ns | 554 (96.8%) | 169 (94.9%) | 98 (93.3%) | ns |
| Remdesivir./Baricitinib, n (%) | 0 | 0 | 0 | na | 11 (1.9%) | 3 (1.6%) | 3 (2.8%) | 0.02 |
| Remdesivir./Dexamethasone, n (%) | 0 | 0 | 0 | na | 4 (0.6%) | 0 | 1 (0.95%) | ns |
| Dexamethasone./Baricitinib, n (%) | 0 | 0 | 0 | na | 1 (0.1%) | 2 (1.1%) | 0 | ns |
| NA | 5 | 13 | 6 |  | 0 | 0 | 2 |  |
| **Previous steroid** |  |  |  |  |  |  |  |  |
| Dexamethasone, n (%) | 0 | 0 | 0 | na | 202 (35.3%) | 66 (37%) | 40 (38%) | ns |
| Prednisone, n (%) | 0 | 0 | 0 | na | 28 (4.8%) | 0 | 5 (4.7%) | 0.001 |
| Betamethasone, n (%) | 0 | 0 | 0 | na | 12 (2%) | 7 (3.9%) | 2 (1.9%) | ns |
| Dexamethasone./Prednisone, n (%) | 0 | 0 | 0 | na | 6 (1.04%) | 2 (1.1%) | 0 | ns |
| Dexamethasone./Betamethasone, n (%) | 0 | 0 | 0 | na | 5 (0.87%) | 4 (2.2%) | 0 | ns |
| NA | 5 | 13 | 6 |  | 0 | 0 | 2 |  |
| **Symptoms** |  |  |  |  |  |  |  |  |
| Fever, n (%) | 19 (48.7%) | 10 (31.2%) | 12 (30%) | 0.01 | 409 (71.5%) | 134 (75.2%) | 57 (54.2%) | <0.001 |
| Cough, n (%) | 19 (48.7%) | 19 (59.3%) | 15 (37.5%) | ns | 333 (58.2%) | 107 (60.1%) | 68 (64.7%) | ns |
| Diarrhea, n (%) | 4 (10.2%) | 1 (3.1%) | 4 (10%) | 0.01 | 61 (10.6%) | 17 (9.5%) | 6 (5.7%) | ns |
| Myalgias, n (%) | 9 (23%) | 7 (21.8%) | 11 (27.5%) | ns | 382 (66.7%) | 111 (62.3%) | 65 (61.9%) | ns |
| Arthralgias, n (%) | 10 (25.6%) | 10 (31.2%) | 8 (20%) | 0.002 | 163 (28.4%) | 47 (26.4%) | 24 (22.8%) | ns |
| Nasal congestion, n (%) | 0 | 0 | 2 (5%) | na | 51 (8.9%) | 21 (11.7%) | 11 (10.4%) | ns |
| Pharyngodynia, n (%) | 7 (17.9%) | 7 (21.8%) | 11 (27.5%) | ns | 140 (24.4%) | 57 (32%) | 29 (27.6%) | 0.05 |
| Anosmia, n (%) | 2 (5.1%) | 3 (9.3%) | 2 (5%) | ns | 43 (7.5%) | 14 (7.8%) | 3 (2.8%) | ns |
| Headache, n (%) | 13 (33.3%) | 7 (21.8%) | 13 (32.5%) | <0.001 | 190 (33.2%) | 60 (33.7%) | 33 (31.4%) | ns |
| NA | 5 | 20 | 14 |  | 0 | 0 | 18 |  |

**COPD:** Chronic obstructive pulmonary disease**NA:** Data not available
P values were obtained from Chi-square or Fisher exact test.
**ns**= non-significant, **na**= not available comparison.

For outpatients, the median age was 39.5 years (IQR: 30-52), 47.7% were female, and 64% of the patients were either partially (28.8%) or fully (36%) vaccinated. The most frequently administered vaccine in fully vaccinated patients was Pfizer (70%), followed by Sinovac (12%) and AstraZeneca (10%). The most commonly administered vaccines in partially vaccinated patients were AstraZeneca (40.6%) and Pfizer (37%). 13.5% of the patients had at least one comorbidity, obesity being the most common (14.4%), particularly in unvaccinated (12.8%) and partially vaccinated (21.8%). For fully vaccinated outpatients, the most common comorbidity was hypertension (12.5%). Only eight patients received dexamethasone (7%) as in-hospital treatment, with the highest prevalence in the unvaccinated group (12.8%). Among the outpatients, unvaccinated patients reported fever (48.7%, Chi-square test, *p* = 0.01) and headache (33.3%, Chi-square test, *p* = 0.01) more frequently than vaccinated individuals. While partially vaccinated patients reported a higher prevalence of arthralgias (31.2%, Chi-square test, *p* = 0.002). Diarrhea was less frequently reported in partially vaccinated patients (3.1%, Chi-square test, *p* = 0.01). The rest of the symptoms were equally reported.

For hospitalized patients, the median age was 52 (IQR: 41-64), and 37% were female. Most patients were unvaccinated (66.9%), while 20.8% were partially vaccinated and 12.2% were fully vaccinated. For fully vaccinated patients, the most frequently administered vaccines were Sinovac (33.3%), Sputnik (20%), and Cansino (17.1%), while for partially vaccinated patients, AstraZeneca (38.7%), Sputnik (28.6%), and Pfizer (19%) were the most frequently administered. 34.6% had at least one comorbidity. Obesity was the most frequent comorbidity in the three compared groups (unvaccinated: 44.5%, partially vaccinated: 46%, and fully vaccinated: 44.7%). In general, fully vaccinated patients were significantly older (median: 70, Chi-square test *p* < 0.001) and showed higher prevalence of diabetes (40%, Chi-square test *p* < 0.001), hypertension (53.3%, Chi-square test *p* < 0.001), and three or more comorbidities (37.1%, Chi-square test *p* < 0.001). Above 90% of hospitalized patients (regardless of the vaccination status) received in-hospital treatment, being dexamethasone the most administered (unvaccinated: 96.8%, partially vaccinated: 94.9%, and fully vaccinated: 93.3%). Fever and pharyngodinia were more frequently reported by partially vaccinated patients (75.2% and 32%, respectively, Chi-square test, *p* < 0.05). The rest of the symptoms were equally reported.

Furthermore, we found no differences in the radiological patterns between study groups. Most of the patients presented a crazy-paving pattern regardless of the vaccination status (unvaccinated: 57.6%, partially vaccinated: 67.4%, fully vaccinated: 68.5%), while the least showed pattern was consolidation (unvaccinated: 6.2%, partially vaccinated: 3.3%, fully vaccinated: 4.8%).

### Impact of vaccination status and age on the severity and clinical parameters of COVID-19 patients

We compared the distribution of COVID-19 severity, severity indexes, O_2_ requirement, and laboratory parameters among patients with different vaccination status and age (Fig. 1-2). Overall, due to the timing of the vaccination campaign in Mexico, most of the vaccinated patients were received from June to August, while unvaccinated patients arrived at the hospital in equal proportion during the sampling months (Suppl. Fig. S1).

**Figure 1.**
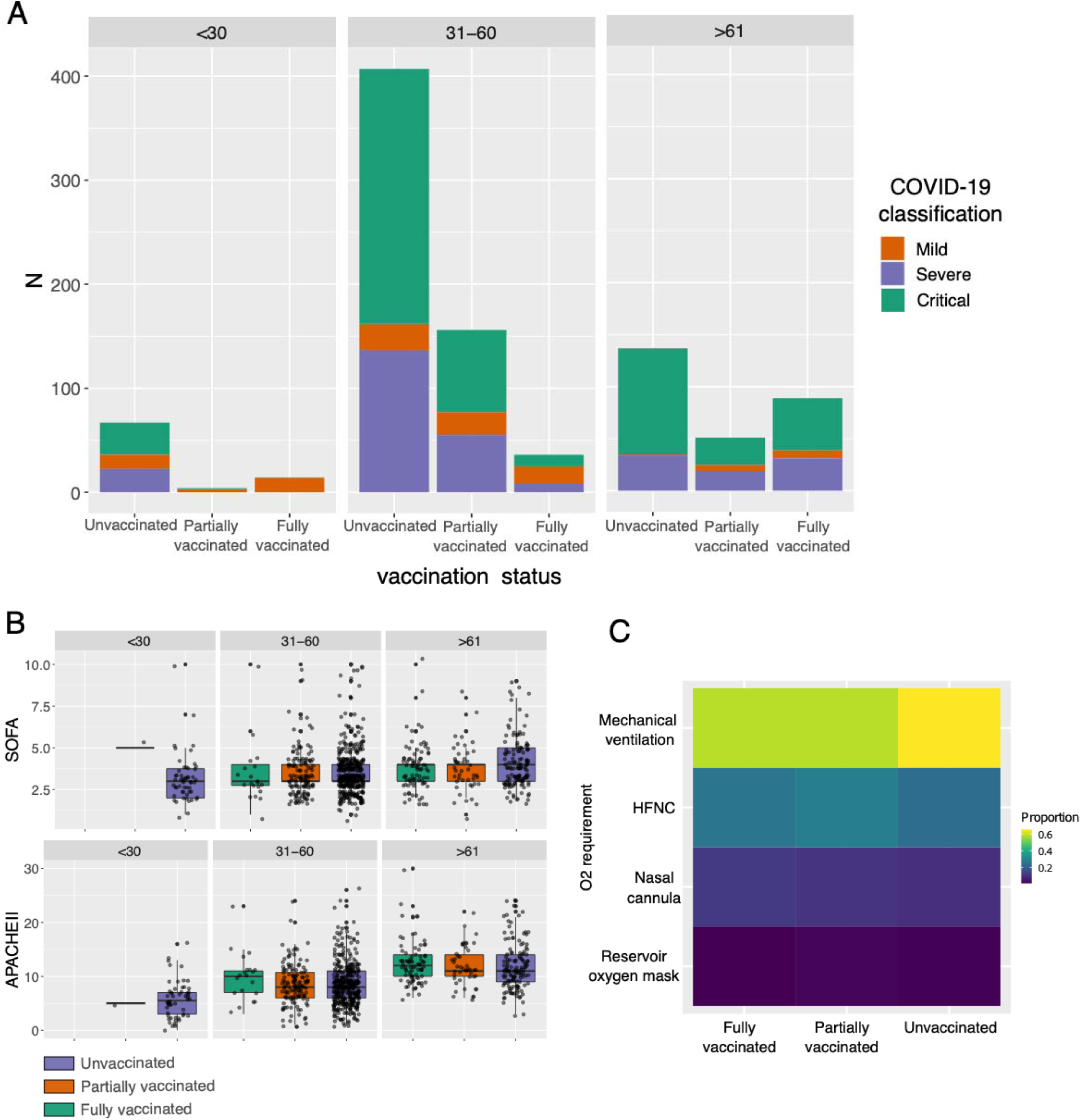
Impact of vaccination status and age in severity and clinical parameters in COVID-19 patients. **A**. Barplot shown the proportion of COVID-19 severity classification among unvaccinated, partially vaccinated, and fully vaccinated patients between age groups. Statistically significant differences are available in Supplementary Table S1. **B**. Boxplot of Sequential Organ Failure Assessment (SOFA) score and Acute Physiology and Chronic Health Evaluation II (APACHE) among patients with different vaccination status and age. **C**. Heatmap of the O_2_ requirement among patients with different vaccination status. HFNC: High Flow Nasal Cannula.

In general, we found that a large proportion of patients were classified either as severe or critical, regardless of age and vaccination status (Fig. 1A). Nonetheless, the highest proportion of patients with mild disease was found in fully vaccinated patients under 30 years old (Suppl. Table S1, Chi-square test *p* < 0.001), while the highest proportion of critical patients were unvaccinated older than 61 years old (Suppl. Table S1, Chi-square test *p* < 0.001). It is essential to highlight that for older patients (>61 years) the vaccination status seems to be associated with less disease severity, reducing the number of critical patients (Suppl. Table S1, unvaccinated against vaccinated patients; Chi-square test *p* = 0.001) and slightly increasing the number of patients with mild disease in the vaccinated groups (Suppl. Table S1, Chi-square test *p* < 0.002). Furthermore, we do not find statistical differences in the most commonly used ICU scales (Fig. 1B) (SOFA and APACHE II). Nonetheless, we detected that, although most of the patients were subjected to mechanical ventilation, the highest proportion of intubated patients was in the unvaccinated group (Fig. 1C).

Finally, we also investigated whether patients with different vaccination status and age differ in laboratory parameters (Fig. 2). The most remarkable differences were found in the unvaccinated group, particularly when comparing patients <30 years with >61 years. For instance, we found that, regardless of the vaccination status, patients older than 61 years exhibited the highest values for blood urea nitrogen (BUN), urea, creatinine (only statistically significant in unvaccinated patients), and D-dimer. In addition, patients in the age groups of <30 and 31-60 showed the highest values for lymphocytes, hematocrit, and albumin (only statistically significant in unvaccinated patients).

**Figure 2.**
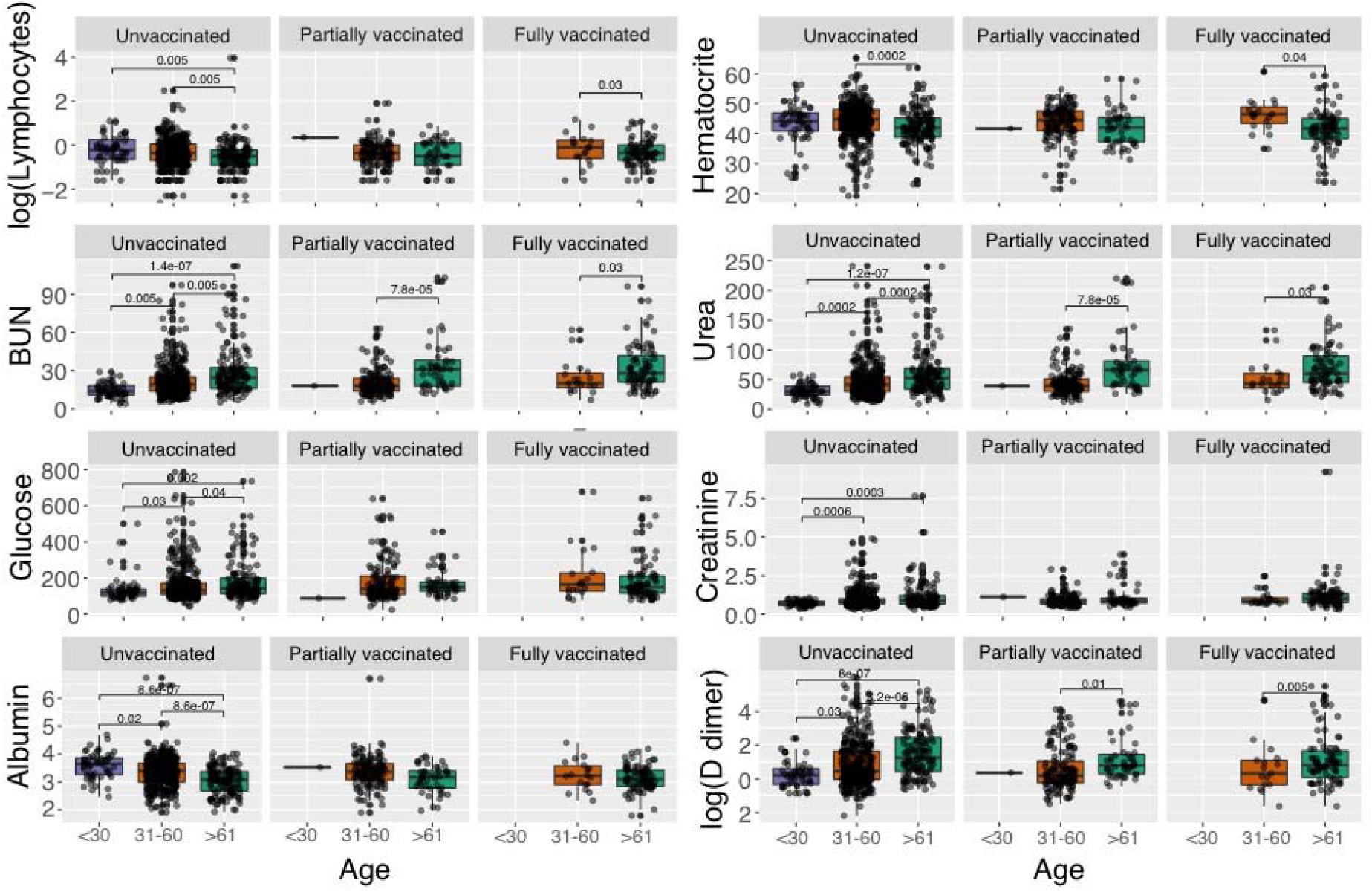
Laboratory parameters that significantly vary among vaccination status and age group. We used logarithmic distribution in the lymphocytes and D dimer parameters for visualization purposes. Statistically significant differences are given by Wilcoxon rank-sum test. Units: Lymphocytes (10^3^/mm^3^), Blood Urea Nitrogen (BUN) (mg/dL), Glucose (mg/dL), Albumin (g/dL), Hematocrit (%), Urea (mg/dL), Creatinine (mg/dL), D dimer (mg/dL).

### Impact of different vaccines on the severity of the disease

By September 2021, more than 100 million doses of seven different vaccines were administered in the Mexican territory, with AstraZeneca (42.9%), Pfizer (32.6%), Sinovac (19.7%), and Sputnik (9.7%) being the most common (Fig. 3A). This heterogeneity of vaccine strategies is also found in our cohort, where 142 patients were fully vaccinated with five different vaccines being Pfizer (29.5%), Sinovac (28.1%), Sputnik (16.1%), AstraZeneca (13.3%), and Cansino (11.9%) the most administered (Fig. 3B). This allowed us to analyze the effect of the different vaccines on the severity of the disease. To isolate the age effect (Suppl. Figure S2), we kept only fully vaccinated patients above 61 years old (N=88) and tested for differences in the severity of the disease, O_2_ requirement, and severity indexes (Fig. 3C-E). Regarding COVID-19 classification, we found that all patients vaccinated with AstraZeneca (N=12) or Cansino (N=7) coursed a severe or critical disease without patients in the mild disease group (Fig. 3C), although this may be due to small sample sizes. Also, we found the highest proportion of patients with mild disease in the Pfizer group (Fig. 3C and Suppl. Table S2) (16.6%, Chi-square test *p* = 0.02). Furthermore, patients vaccinated with AstraZeneca, Sinovac, and Sputnik had mostly a critical disease (66%, 51.4%, and 59%, respectively). Regarding O_2_ requirement, although most of the patients were subjected to mechanical ventilation, AstraZeneca was the group with the highest proportion (66%). Pfizer was the group with the highest proportion of patients with a nasal cannula (16.6%, Chi-square test *p* = 0.01), and the Cansino group had the highest proportion of high flow nasal cannula (HFNC) (42.8%, Chi-square test *p* = 0.01). Finally, we found that the Sinovac group had significantly higher values for SOFA (Fig.3E) compared to Pfizer (Wilcoxon rank-sum test, *p* = 0.01) and AstraZeneca (Wilcoxon rank-sum test, *p* = 0.02). We did not find statistically significant differences in the APACHE II severity index.

**Figure 3.**
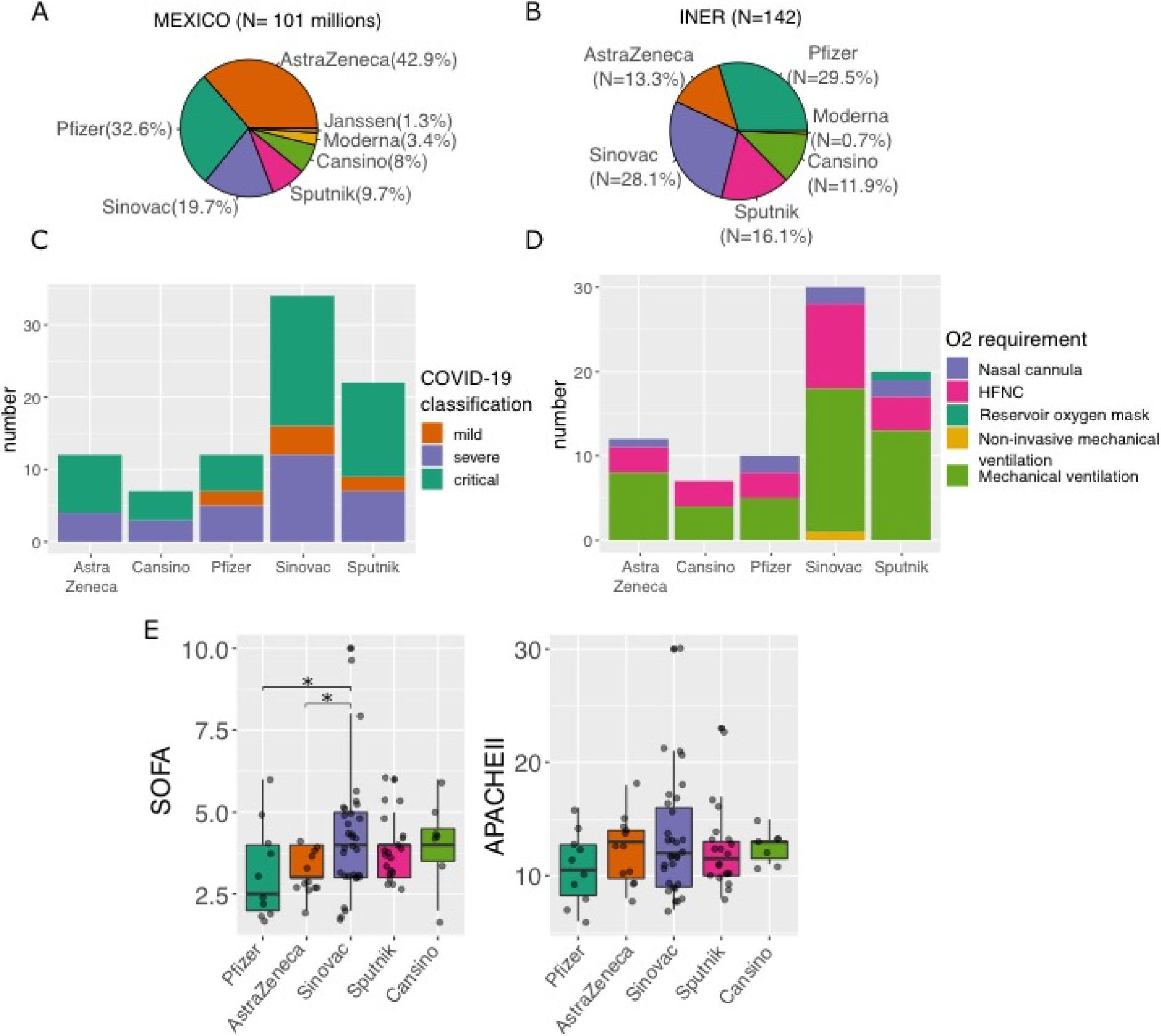
Distribution of vaccine strategies and their impact on disease severity in fully vaccinated patients. **A**. Piechart of the proportion of the vaccines applied at the national level. **B**. Piechart of the proportion of the different vaccines applied at the INER. **C**. Barplot shown the proportion of COVID-19 classification in patients with different vaccines. **D**. Barplot depicting the proportion of O_2_ requirement in patients with different vaccines. **E**. Boxplot of Sequential Organ Failure Assessment (SOFA) score and Acute Physiology and Chronic Health Evaluation II (APACHE) among patients with different vaccines. Statistically significant differences are available in Supplementary Table S2-3. In C-E panels only patients >61 years old were included.

### Clinical factors affecting COVID-19 clinical outcome

Of the hospitalized patients, 77.1% were discharged fully recovered, while 21.2% died due to complications (Table 2), with the highest proportion of deceased patients in the unvaccinated group (70.8%). Regarding severity indexes, fully vaccinated patients had the highest median of APACHE II (median: 11, Wilcoxon rank-sum test, *p* < 0.001). Most of the patients were subjected to supplemental oxygen, with mechanical ventilation (unvaccinated: 65.5%, partially vaccinated: 58.9%, fully vaccinated: 58%), HFNC (unvaccinated: 23.6%, partially vaccinated: 28.4%, fully vaccinated: 26.6%), and nasal cannula (unvaccinated: 9.4%, partially vaccinated: 10.6%, fully vaccinated: 11.4%) being the most common.

**Table 2.** Outcome, severity indexes, and O_2_ requirement among hospitalized patients with different vaccination status.

|  | Hospitalized (N=855) |  |  | P value |
| --- | --- | --- | --- | --- |
|  | Unvaccinated<br>(N=572)<br>[66.9%] | Partially<br>vaccinated<br>(N=178) [20.8%] | Fully<br>vaccinated<br>(N=105) [12.2%] |  |
| **Outcome** |  |  |  |  |
| Recovered, n (%) | 433 (71.6%) | 139 (78%) | 88 (83.8%) | **0.05** |
| Deceased, n (%) | 129 (22.5%) | 37 (20.7%) | 16 (15.2%) | *ns* |
| NA | 10 | 2 | 1 |  |
| **Severity indexes** |  |  |  |  |
| APACHE II, med(IQR) | 8.5 (6-11) | 9 (6.2-11) | 11 (9-14) | **<0.001** |
| SOFA, med(IQR) | 3 (3-4) | 3 (3-4) | 4 (3-4) | *ns* |
| GLASGOW, med(IQR) | 15 (15-15) | 15 (15-15) | 15 (15-15) | *ns* |
| **O<sub>2</sub> requirement** |  |  |  |  |
| Nasal cannula, n (%) | 54 (9.4%) | 19 (10.6%) | 12 (11.4%) | *ns* |
| HFNC, n (%) | 135 (23.6%) | 50 (28%) | 28 (26.6%) | *ns* |
| Mechanical ventilation, n (%) | 375 (65.5%) | 105 (58.9%) | 61 (58%) | *ns* |
| Reservoir oxygen mask, n (%) | 7 (1.2%) | 3 (1.6%) | 1 (0.9%) | *ns* |
| Non-invasive mechanical ventilation, n (%) | 1 (0.1%) | 1 (0.5%) | 1 (0.9%) | *ns* |
| NA | 0 | 0 | 2 |  |

**NA:** Data not available
P values were obtained from Chi-square or Fisher exact test.
*ns*= non-significant, *na*= not available comparison.

Moreover, we applied multivariate statistical approaches to determine the factors that could explain mortality in COVID-19 patients. From the two fitted Generalized Linear Model (GLM) (Fig. 4A), we found that the presence of immune disease (OR: 3.12, 95% CI: 1.09-8.34, *p* = 0.02), age above 61 years old (OR: 3.51, 95% CI: 2.3-5.2, *p* = 5.9e-10), platelets count under 175 ×10^9^/l (OR: 1.49, 95% CI: 0.98-2.2, *p* = 0.05), lactate dehydrogenase (LDH) > 600 u/L (OR: 1.89, 95% CI: 1.2-2.7, *p* = 0.001), and D dimer > 1.8 ug/mL (OR: 1.45, 95% CI: 0.98-2.1, *p* = 0.05) were factors associated with higher probability of death. While been fully vaccinated was associated with improved outcomes (OR: 0.25, 95% CI: 0.12-0.46, *p* = 2.89e-05). Odds ratios and estimates for all variables in the model are available in Supplementary Table S4. Finally, age was also found to correlate with mortality (Cox test, *p* < 0.001) in the Kaplan-Meier curves (Fig. 3C), as well as D-dimer above 1.87 µg/mL independently from vaccination or comorbidities (Cox test, *p* = 0.02).

**Figure 4.**
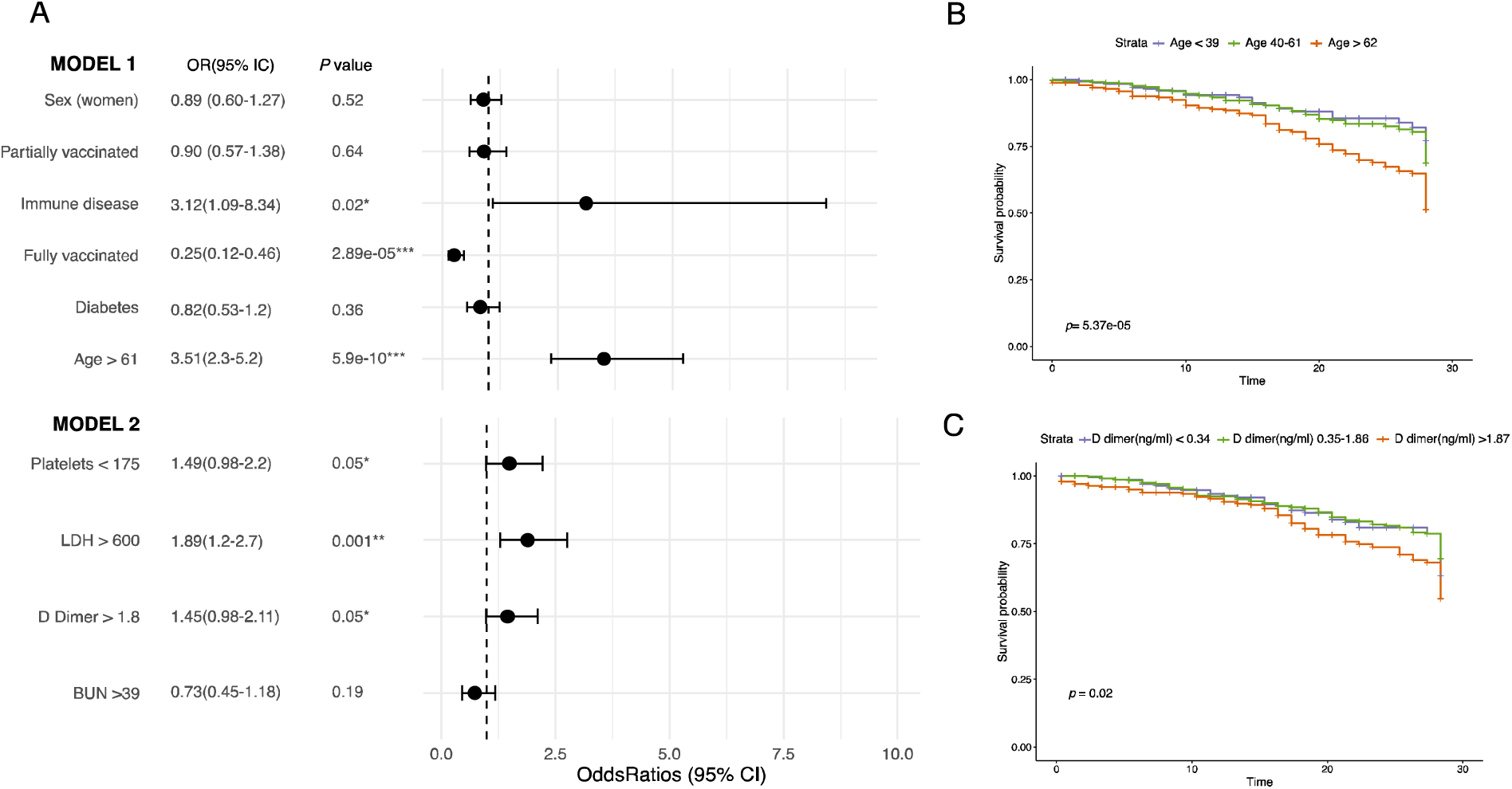
Variables affecting outcome in COVID-19 patients. **A**. Forest plot of the two fitted Generalized Linear Model (GLM) representing the odds ratio and 95% IC for each variable. Estimates are available in Supplementary Table S4. **B**. Kaplan-Meier curve depicting the survival probability of patients with different age ranges. **C**. Kaplan-Meier curve depicting the survival probability of patients with different values for D dimer. All models were constructed using outcome (deceased or recovered) standardized at 28 days. Units: Platelets (×10^9^/l), Lactate dehydrogenase (LDH) (u/L), D dimer (ug/dL), and Blood Urea Nitrogen (BUN) (mg/dL).

### SARS-CoV-2 lineages: phylogeny, distribution, and its impact on disease severity

We identified the lineages of 386 (38%) samples by partial or complete genome sequencing (Table 3). From this subset of samples, we detected VOC such as Alpha (2.3%), Gamma (7.2%), Delta (68.6%), and Mu (3.6%), originally designated as VOI. We also detected sequences belonging to lineage B.1.1.519 (14.7%) and other lineages (2.86%). The distribution of lineages with a clear dominance of Delta, and in less proportion of B.1.1.519 was also observed between March and September 2021 nationwide, as well as in the State of Mexico, and Mexico City (Fig.5A, statistical differences are available in Supplementary Table S5).

**Table 3.** SARS-CoV-2 lineages between outpatients and hospitalized patients.

| Lineages | Outpatients (N=123) | Hospitalized (N=263) | P value |
| --- | --- | --- | --- |
| Alpha, n (%) | 2 (1.5%) | 7 (2.6%) | ns |
| Gamma, n (%) | 4 (3.1%) | 24 (9.1%) | ns |
| Delta, n (%) | 104 (82.5%) | 163 (61.9%) | **<0.001** |
| Mu, n (%) | 3 (2.3%) | 11 (4.1%) | ns |
| B.1.1.519, n (%) | 7 (5.5%) | 50 (19%) | **0.001** |
| Other, n (%) | 3 (2.3%) | 8 (3.04%) | ns |

P values were obtained from Chi-square or Fisher exact test.

**Figure 5.**
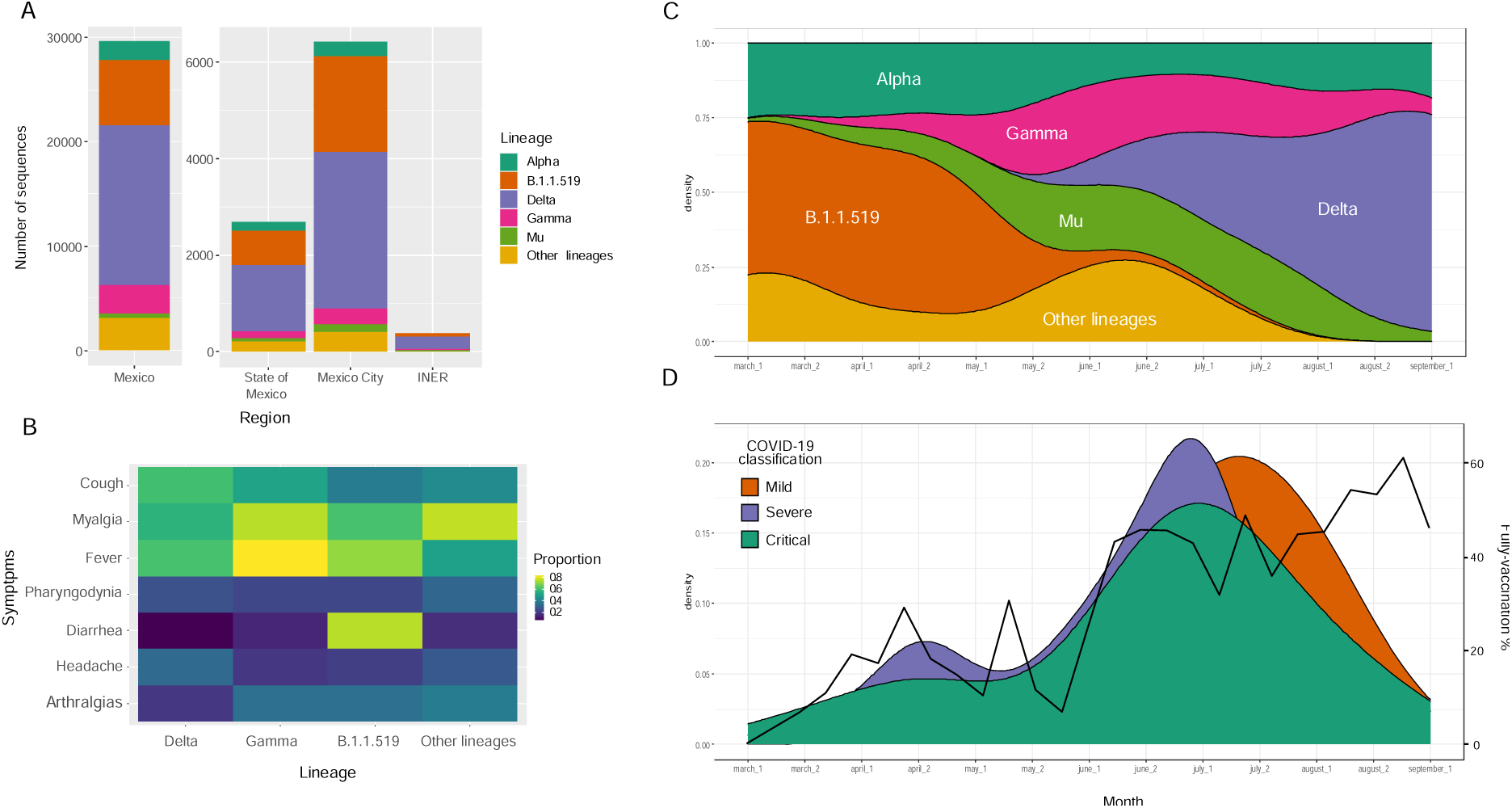
SARS-CoV-2 lineages distribution and its impact on disease severity. **A**. Distribution of SARS-CoV-2 lineages at the country level (Mexico), Metropolitan area (the State of Mexico and Mexico City), and our research center (INER). **B**. Heatmap of the principal symptoms among patients infected with the different SARS-CoV-2 lineages. **C**. Dominance of lineages in our cohort along time (months). **D**. Distribution of patients with either mild, severe, or critical COVID-19 classification and percentage of fully vaccinated patients in our cohort through time (months).

Furthermore, we also found differences in the distribution of SARS-CoV-2 lineages between outpatients and hospitalized patients (Table 3). In particular, we found the highest proportion of Delta variant in outpatients (82.5%, Chi-square test, *p* < 0.001), while B.1.1.519 represented the highest proportion in hospitalized patients (19%, Chi-square test, *p* = 0.001). Notably, the SARS-CoV-2 variant distribution displayed temporal variations, with B.1.1.519 being more prevalent in March and April 2020, and Delta becoming the predominant lineage in July (Fig. 5C). Furthermore, we found a general increase in the proportion of vaccinated patients during the study period (Fig. 5D). These temporal variations make complex the analysis of potential associations between disease severity and lineages. It is worth mentioning that the number of critical and severe patients (green and purple colors) seems to decrease as the vaccination increases (Fig. 5D). Altogether, we found no significant association between disease severity and lineages (Supplementary Fig. S3). Nonetheless, when analyzing the symptoms associated with patients infected with different lineages, we noted that diarrhea tended to be associated more frequently with B.1.1.519 infections (Fig. 5B).

Finally, the phylogenetic analyses based on complete Spike sequences showed, as expected, a specific clustering by lineages, with the Delta variant representing 69% of the samples (Suppl. Figure S4). The sequences of patients with different vaccination statuses or severity classifications did not form specific clusters; suggesting no association between SARS-CoV-2 lineages and vaccination status or COVID-19 severity.

## DISCUSSION

Vaccines have significantly improved the COVID-19 survival rate around the world, but they are less effective at protecting from SARS-CoV-2 infections, leading to mostly mild infections in vaccinated individuals. Understanding the impact of viral variants, and host factors on disease severity is critical to guide COVID-19 vaccination campaigns and the implementation of protective measures. In this retrospective study, we analyzed the severity of COVID-19 infections in a group of individuals that had been vaccinated with all different types of vaccines applied to the Mexican population. We compared this group with unvaccinated individuals and performed additional analyses, including virological features obtained from genomic surveillance.

### Impact of host factors and vaccination status on COVID-19 severity

It is known that host factors such as advanced age and the presence of comorbidities significantly worsen clinical outcomes for COVID-19 patients [28]. The Mexican population is characterized by a high prevalence of comorbidities [29], which is reflected in our cohort, with diabetes (20.5%), hypertension (25.5%), and obesity (39.4%) being the most common. Moreover, it is important to consider that the vaccination campaign in Mexico started at the end of 2020 and first focused on health care workers before moving on to individuals over 60, 50, and 40 years old in February, May, and June, respectively. As a result, in our cohort, the prevalence of comorbidities was even higher in fully vaccinated patients (Table 1), with more than 90% presenting at least one comorbidity, and over 75% were older than 60 years of age. Some studies have shown that the presence of comorbidities and advanced ages can also affect vaccines’ effectiveness [12]. Moreover, in both COVID-19 [6,22,30] and Influenza disease [31], it has been shown that advanced age is related to a lower immune response to vaccination. Furthermore, a recent study showed that although vaccinated patients were less likely to present severe disease, vaccinated patients with comorbidities and older age developed severe disease [6]. This is similar to our study where we observed that fully vaccinated patients presented the highest values of severity on the APACHE II scale (Table 2). Nonetheless, when compared to unvaccinated individuals, the vaccinated group showed the lowest mortality rate (Table 2) and required mechanical ventilation less frequently (Fig.1C). These observations confirm that although the vaccine’s protective effect may be reduced in individuals with risk factors, they still provide protection against fatal outcomes.

Moreover, we found that a significant proportion of patients had either severe or critical outcomes, regardless of age and vaccination status (Fig.1A). Although this could be a consequence of studying a cohort of patients in a tertiary hospital limited to the attention of critically ill individuals, it also could be the result of the presence of comorbidities and host genetic factors [32,33]. However, it is essential to highlight that the highest proportion of patients with mild disease was in those fully vaccinated and under 30 years of age (Suppl. Table S1), this evidences the combined protective effect of younger ages and complete vaccination schemes [34].

In addition, we analyzed laboratory parameters by vaccination status and age groups. Regardless of the vaccination status, patients older than 61 years exhibited the highest values for BUN, urea, creatinine, and D-dimer (Fig. 2). This is of special relevance since such parameters are known as markers of disease severity and are commonly found elevated in individuals with acute infections [35]. Moreover, urea, BUN, and creatinine are also markers for renal damage and acute kidney injury (AKI), which are conditions that strongly impact mortality [36]. Also, hypoalbuminemia, found in older COVID-19 patients, could be a manifestation of a pro-inflammatory state [37]. Finally, high levels of D-dimer have been found related to mortality in patients with infection or sepsis [38].

### Impact of different vaccines on the severity of the disease

It is important to point out that this study was not designed to assess vaccine effectiveness against COVID-19. However, Mexico is one of the few countries that used seven different vaccines, offering a unique opportunity to compare these vaccine types and to look for differences in severity and/or mortality. In this study, by analyzing solely patients above 61 years old we found, in general, a high proportion of critical patients but low mortality rates regardless of the vaccine applied (Suppl. Table S2). Nonetheless, our findings of lower SOFA index, a higher proportion of patients with mild disease, and higher usage of nasal cannula in patients vaccinated with the Pfizer vaccine may suggest a lower severity of disease in the group that received this vaccine. Some studies have shown that the vaccination efficacy of mRNA vaccines does not decrease in individuals with comorbidities [39]. Still, no causal associations can be made from this finding, and more investigations are needed to confirm if there is a difference in severity associated with the vaccines and what are the factors behind it.

### SARS-CoV-2 lineages and their impact on disease severity

Understanding the impact of SARS-CoV-2 lineages on disease severity in both unvaccinated and vaccinated individuals is of major importance, especially since such variants could still emerge due to the evolution pressure driven by the population immunity [5,40,41]. As part of our genomic surveillance program, we characterized the circulating SARS-CoV-2 lineages at the time of the study and analyzed the disease severity and clinical presentation associated with infections caused by particular lineages. Alpha and B.1.1.519 were the predominant lineages from March to May 2021, with Delta becoming the most prevalent by July 2021. In addition, other variants such as Gamma and Mu were also detected, mainly between May and July 2021(Fig. 5-6).

Furthermore, we detected a tendency in patients >60 years old infected with either Delta or B.1.1.519 lineages to course a more severe disease (Suppl. Figure S3). Although this finding was not statistically significant in our study, other studies have reported more severe disease in infections associated with these lineages [10,42]. In particular, mutations in the spike (S) protein of the virus which confer a higher affinity of the S1 domain to the angiotensin converter enzyme 2 (ACE2) are one of the main reasons behind the higher pathogenicity of the Delta variant [42].

Regarding symptomatology, there were no remarkable differences except for infections with the B.1.1.519 lineage, since patients reported diarrhea more frequently (Fig.5B). This contrasts with other studies where they found that fever, dyspnea, and sore throat were more prevalent in patients infected with Delta [6,43]. However, a previous study found that dyspnea, cyanosis, and particularly diarrhea were the most frequent symptoms in patients infected with the B.1.1.519 lineage in Mexico [10].

Moreover, previous studies analyzing large datasets have found differences in disease severity among patients infected with different lineages [44,45]. Nonetheless, studies with small datasets have failed in detecting such differences [46,47]. Altogether, the reduced sample size and the temporal variation of the lineages may be affecting our results and hinder comparative analyses between lineages and vaccination status or disease severity.

### Clinical factors affecting COVID-19 clinical outcome

We determined the main factors that lead to fatal outcomes in our cohort. According to the fitted Generalized Linear Models (GLM) (Fig. 4), full vaccination was found to be a protective factor against death by COVID-19. On the contrary, platelets <175 ×10^9^/l, age above 61 years old, immune disease, D-dimer > 1.8 ug/mL, and lactate dehydrogenase (LDH) above 600 u/L were found as risk factors. Notably, thrombocytopenia and high levels of LDH have been found associated with severe COVID-19 in other studies [48,49]. Furthermore, contrary to other studies [50] we did identify advanced age as a risk factor itself in COVID-19 patients. It is known that the decline of physiologic functions and the immunosenescence associated with increasing age can negatively affect the host response to infectious diseases [6,22,30]. Finally, we hypothesize that complete vaccination may diminish age negative effect since we found that despite that most of the fully vaccinated patients were >61 years old (78%), 83.3% were discharged from the hospital.

Our study includes a considerable number of critical and severe patients enrolled, which allowed us to analyze the impact of vaccination, vaccine type, SARS-CoV-2 lineages, and several host factors on mortality and clinical presentation. However, the source is a single referral hospital, and a pandemic wave mostly by the Delta variant, preventing an analysis among vaccination status and lineages.

## CONCLUSIONS

Comorbidities and advanced ages were the main risk factors, and complete vaccination schemes with different vaccine types (Pfizer, AstraZeneca, Cansino, SinoVac, and Sputnik) were the most significant protective factor against death by COVID-19. Moreover, we did not find strong associations between a particular lineage (Alpha, Gamma, B.1.1.519, and Delta) and disease severity, highlighting the predominant role of host factors such as age, comorbidities, and vaccination status on COVID-19 outcome.

Complete vaccination schemes for the whole population, are key to reducing both hospitalization and death. Nevertheless, public health policy should also focus on the control of comorbidities in the long term, which will improve clinical outcomes not only for COVID-19 but also for other current and emerging diseases. Genomic surveillance is essential to identify circulating VOC, which can help to guide the implementation of targeted epidemiological control measures and laboratory characterization for diagnostic and research purposes.

## Data Availability

The genomic information generated during the current study is available in the GenBank repository, accession numbers: ON158371-ON158443.
The clinical datasets used during the current study are available from the corresponding author on reasonable request.

## Acknowledgments

We thank Eduardo Márquez García from Unidad de Biología Molecular, INER for technical assistance in Illumina sequencing. We also thank all physicians of the ICU for assistance with patient management.

## Authors contributions

Conceptualization: PGD, VAT, SAR, RPP, CFA, and JAVP. Methodology: CB, KTD, MMF, MPG, MEJC, FMN, and JAVP. Formal Analysis: AHT, MVR, RSM, and JAVP. Visualization: AHT. Investigation: PGD, MVR, CB, BT, EP, VAT, KTD, MMF, RSM, FJH, EBV, OB, JAMO, and JAVP. Resources: PGD, VAT, SAR, RPP, CFA, and JAVP. Data Curation: AHT. Writing-Original Draft Preparation: AHT, PGD, MVR, CB, VAT, RSM, RPP, CFA, and JAVP. Writing-Review & Editing: All authors. Supervision: CFA, and JAVP. Funding Acquisition: CFA, and JAVP.

## Institutional Review Board Statement

This study was reviewed and approved by the Science, Biosecurity, and Bioethics Committee of the Instituto Nacional de Enfermedades Respiratorias (protocol number B-10-20).

## Informed Consent Statement

Informed consent was provided according to the Declaration of Helsinki. Written informed consent has been obtained from the patients and/or from their relatives or authorized legal guardians to publish this paper.

## Data Availability Statement

The genomic information generated during the current study is available in the GenBank repository, accession numbers: ON158371-ON158443. The clinical datasets used during the current study are available from the corresponding author on reasonable request.

