## Supplementary Material for "Clinical and Virological Features of patients hospitalized with different types of COVID-19 vaccination in Mexico City"

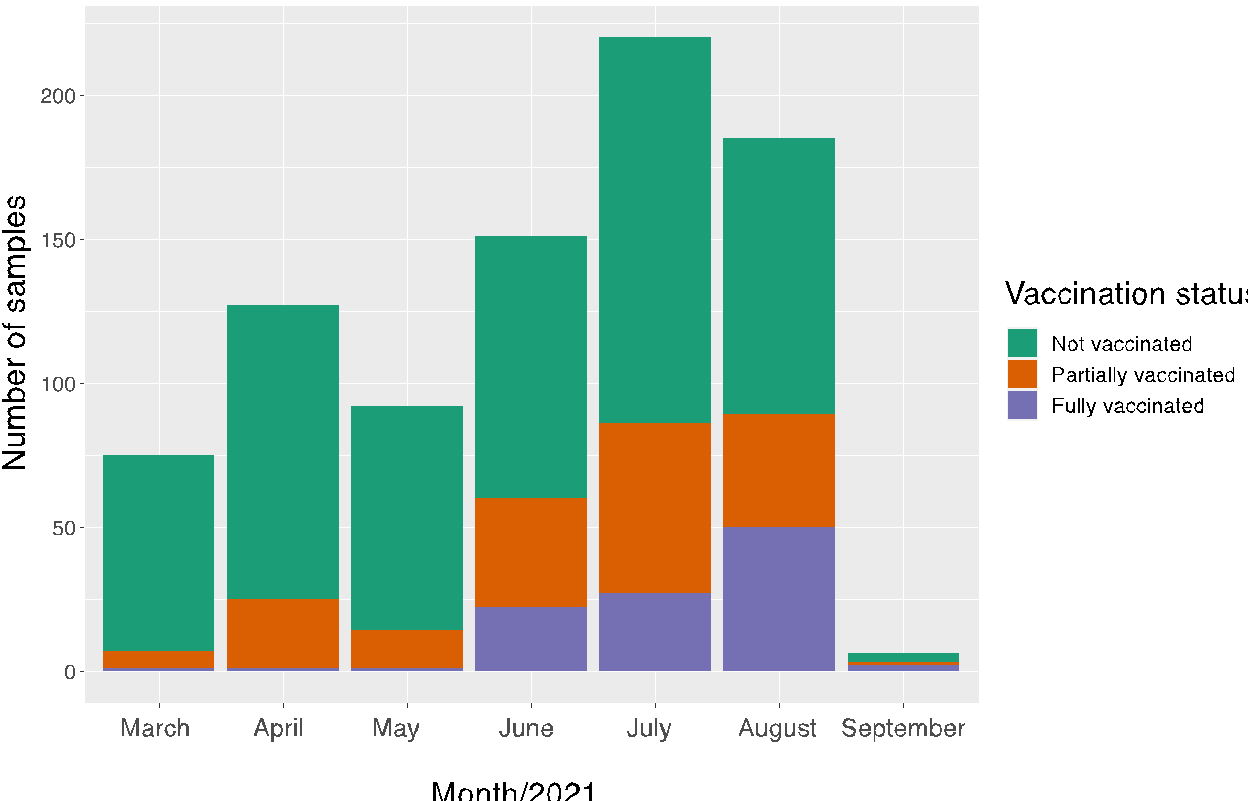

**Supplementary Figure S1. Vaccination status of all patients studied from March to**

**September 2021.**

**Supplementary Table S1. COVID-19 classification among patients with different vaccination status.**

|  | **<30 years old**(N=85) | | | | **31-60 years old**(N=599) | | | | **>61 years old**(N=278) | | | |
| --- | --- | --- | --- | --- | --- | --- | --- | --- | --- | --- | --- | --- |
|  | **NV** | **PV** | **FV** | *p* | **NV** | **PV** | **FV** | *p* | **NV** | **PV** | **FV** | *p* |
| **Mild** | 13(19%) | 3(75%) | 14 (100%) | ***<0.001*** | 25(6.1%) | 22(14.1%) | 17(47.2%) | ***<0.001*** | 1(0.72%) | 6(11.7%) | 8(8.8%) | ***0.002*** |
| **Severe** | 23(34.3%) | 0 | 0 | *ns* | 137(33.6%) | 55(35.2%) | 8(22.2%) | *ns* | 34(24.8%) | 19(37.2%) | 31(34%) | *ns* |
| **Critical** | 31(46.2%) | 1(25%) | 0 | *ns* | 245(60.1%) | 79(50.6%) | 11(30.5%) | ***<0.001*** | 102(74.4%) | 26(50.9%) | 50(55%) | ***0.001*** |
| Total | 67 | 4 | 14 |  | 407 | 156 | 36 |  | 137 | 51 | 90 |  |

NV: not vaccinated

PV: partially vaccinated

FV: fully vaccinated

*P* values were obtained from a Chi-square or Fisher exact test. ns: non-significant. Values statistically significant are in bold.

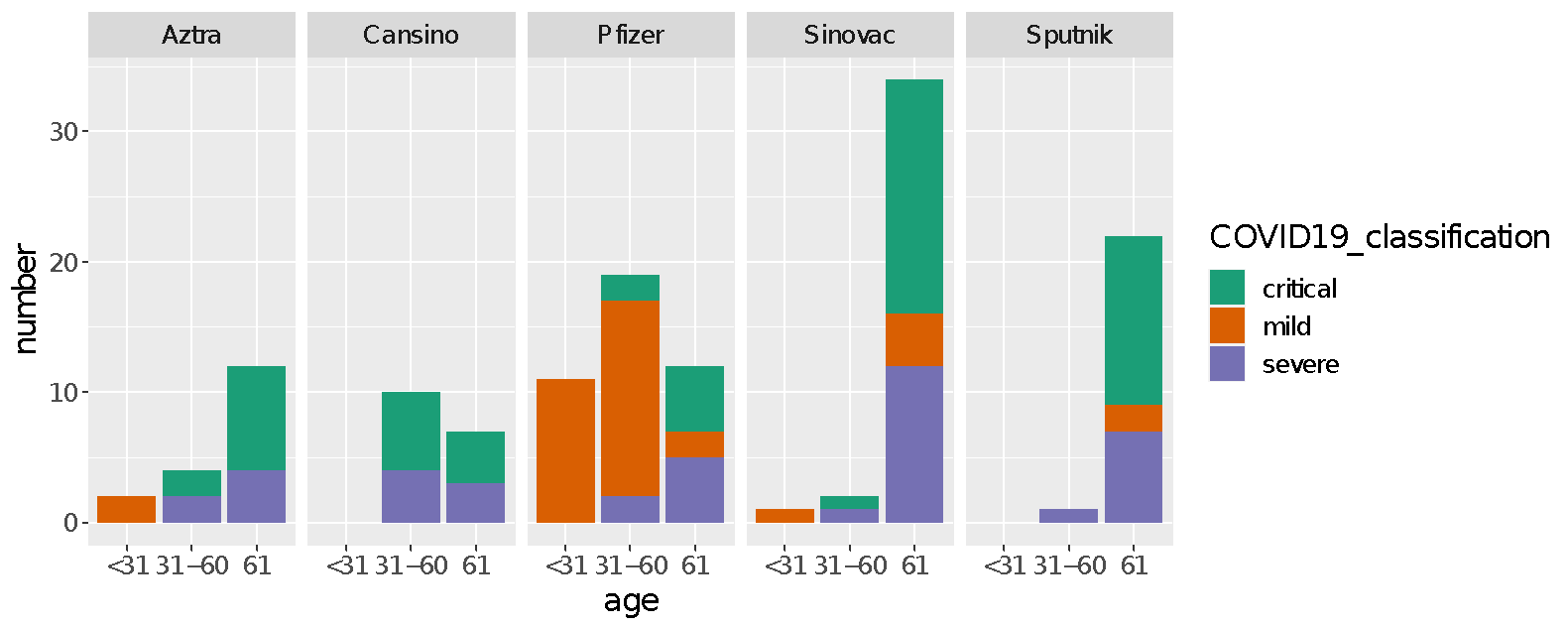

**Supplementary Figure S2. Distribution of ages and severity of the disease in fully vaccinated patients with different vaccine strategies.**

**Supplementary Table S2. Statistical differences in the severity of the disease between patients with different vaccine types and age groups**

|  | **Classification** | **Pfizer** (N=12) | **AstraZeneca** (N=12) | **Sinovac** (N=35) | **Sputnik** (N=22) | **Cansino** (N=7) |
| --- | --- | --- | --- | --- | --- | --- |
| **>61**  (N=88) | Mild | 2(16.6%) | 0 | 4 (11.4%) | 2 (9%) | 0 |
|  | Severe | 5(41.6%) | 4 (33.3%) | 12 (34.2%) | 7 (31.8%) | 3 (42.8%) |
|  | Critical | 5(41.6%) | 8 (66.6%) | 18 (51.4%) | 13 (59%) | 4 (57.1%) |
|  | NA | 0 | 0 | 1 (2.8) | 0 | 0 |
|  | ***P value*** | *ns* | ***0.002*** | ***0.001*** | ***0.002*** | *ns* |

*****We excluded from the table two patients, one vaccinated with Moderna and the other vaccinated with J&J.

*P* values were obtained from a Chi-square or Fisher exact test.

**ns:** non-significant, **na:** comparison not available.

Values statistically significant are in bold.

**Supplementary Table S3. Statistical differences of the O2 requirement between patients with different vaccine type and age group.**

|  | **Classification** | **Pfizer** (N=42) | **AztraZeneca** (N=19) | **Sinovac** (N=40) | **Sputnik** (N=23) | **Cansino** (N=17) | **P *value*** |
| --- | --- | --- | --- | --- | --- | --- | --- |
| **All**  (N= 141)* | Nasal cannula | 4(9.5%) | 1(5.26%) | 2(5%) | 3(13%) | 2(11.7%) | *ns* |
|  | HFNC | 3(7.14%) | 3(15.78%) | 12(30%) | 4(17.3%) | 5(29.4%) | ***<0.001*** |
|  | Reservoir OM | 0 | 0 | 0 | 1(4.34%) | 0 | *na* |
|  | NIMV | 0 | 0 | 1(2.5%) | 0 | 0 | *na* |
|  | MV | 7(16.6%) | 10(52.6%) | 19(47.5%) | 13(56.5%) | 10(58.8%) | ***0.02*** |
|  | None | 0 | 0 | 0 | 0 | 0 | *na* |
|  | NA | 28(66.6%) | 5(26.3%) | 6(15%) | 2(8.6%) |  | ***<0.001*** |
|  | ***P value*** | *ns* | ***0.002*** | ***<0.001*** | ***<0.001*** | *ns* | *na* |
| **>61**  (N=88) | Nasal cannula | 2(16.6%) | 1(8.3%) | 2(5.7%) | 2 (9%) | 0 | ***0.01*** |
|  | HFNC | 3(25%) | 3 (25%) | 10(28.5%) | 4 (18.1%) | 3 (42.8%) | ***0.01*** |
|  | Reservoir OM | 0 | 0 | 0 | 1(4.5%) | 0 | *na* |
|  | NIMV | 0 | 0 | 1(2.8%) | 0 | 0 | *na* |
|  | MV | 5(41.6%) | 8 (66.6%) | 17(48.5%) | 13 (59%) | 4 (57.1%) | *ns* |
|  | None | 0 | 0 | 0 | 0 | 0 | *na* |
|  | NA | 2(16.6%) | 0 | 0 | 0 | 0 | *na* |
|  | ***P value*** | *ns* | ***0.01*** | ***<0.001*** | ***<0.001*** | *ns* | *na* |

*****We excluded from the table two patients, one vaccinated with Moderna and the other vaccinated with J&J.

*P* values were obtained from a Chi-square or Fisher exact test.

**ns:** non-significant, **na:** comparison not available.

Values statistically significant are in bold.

HFNC: high flow nasal cannula.

Reservoir OM: reservoir oxygen mask

NIMV: non-invasive mechanical ventilation

**Supplementary Table S4. Odds ratio and estimates of the two Generalized Linear Models (GLM).**

|  | **Estimate** | **Std. Error** | **Z value** | **Odds ratio** | **95% CI** | **Pr(>z)** |
| --- | --- | --- | --- | --- | --- | --- |
| Model 1 |  |  |  |  |  |  |
| **(Intercept)** | -1.5617 | 0.1375 | -11.35 | - | 0.15-0.273 | ***<0.001****** |
| **Partially vaccinated** | -0.1009 | 0.2223 | -0.454 | 0.90 | 0.57-1.38 | ***0.64*** |
| **Fully vaccinated** | -1.3948 | 0.3335 | -4.182 | 0.25 | 0.12-0.46 | ***2.89e-05****** |
| **Age > 61** | 1.2569 | 0.2030 | 6.192 | 3.51 | 2.31-5.2 | ***5.9e-10****** |
| **Diabetes** | -0.1981 | 0.2171 | -0.912 | 0.82 | 0.52-1.2 | *0.36* |
| **Immune disease** | 1.1385 | 0.5097 | 2.234 | 3.12 | 1.09-8.3 | ***0.02**** |
| **Sex (Women)** | -0.1212 | 0.1884 | -0.643 | 0.89 | 0.60-1.27 | *0.52* |
| Model 2 |  |  |  |  |  |  |
| **(Intercept)** | -1.6283 | 0.2671 | -6.096 | - | 0.11-0.327 | ***1.09e-09****** |
| **Platelets < 175** | 0.3979 | 0.2082 | 1.911 | 1.49 | 0.98-2.22 | ***0.05**** |
| **BUN > 39** | -0.311 | 0.2422 | -1.284 | 0.73 | 0.45-1.18 | ***0.19*** |
| **D Dimer > 1.8** | 0.3693 | 0.1963 | 1.882 | 1.45 | 0.98-2.11 | ***0.05**** |
| **LDH > 600** | 0.6344 | 0.1860 | 3.9 | 1.89 | 1.28-2.75 | ***0.001***** |

**Supplementary Table S5. Statistical differences of the distribution of lineages in the samples sequenced of Mexico, State of Mexico, Mexico City, and INER.**

| **SARS-CoV-2 lineage** | **Mexico** | **State of Mexico** | **Mexico City** | **INER** | ***P* value** |
| --- | --- | --- | --- | --- | --- |
| **Delta**, n (%) | 15,301 (51.6%) | 1,377 (51.2%) | 3248 (50.6%) | 265 (69%) | ***<0.001*** |
| **Alpha**, n (%) | 1,787 (6.03%) | 178 (6.6%) | 291 (4.5%) | 9 (2.3%) | ***<0.001*** |
| **Gamma**, n (%) | 2,749 (9.2%) | 144 (5.36%) | 325 (5%) | 28 (7.2%) | ***<0.001*** |
| **Mu**, n (%) | 434 (1.4%) | 58 (2.1%) | 152 (2.36%) | 14 (3.6%) | ***<0.001*** |
| **B.1.1.519**, n (%) | 6,265 (21.1%) | 711 (26.4) | 1,986 (30.9%) | 57 (14.8%) | ***<0.001*** |
| **Other lineages**, n (%) | 3,085 (10.4%) | 217 (8%) | 412 (6.4%) | 11 (2.8%) | ***<0.001*** |
| Total sequences | 29,621 | 2685 | 6414 | 384 |  |

Data from Mexico, State of Mexico, and Mexico City was taken from gisaid.com.

*P* values were obtained from a Chi-square analysis. Values statistically significant are in bold.

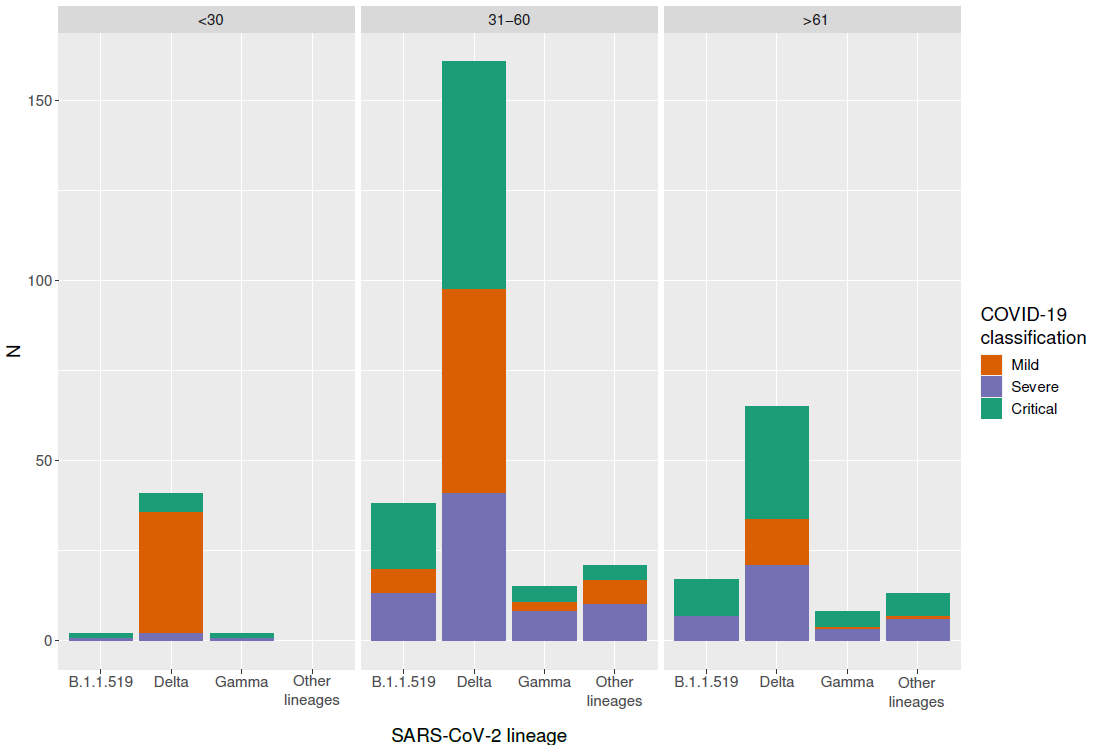

|  | **<30** (N=44) | | | | **31-60** (N=237) | | | | | **>61** (N=103) | | | | |
| --- | --- | --- | --- | --- | --- | --- | --- | --- | --- | --- | --- | --- | --- | --- |
| Status/variant | **Delta** (N=40) | **Gamma** (N=2) | **B.1.1.519**  (N=2) | *p* | **Delta** (N=160) | **Gamma** (N=18) | **B.1.1.519** (N=38) | **Other** (N=21) | *p* | **Delta** (N=65) | **Gamma** (N=8) | **B.1.1.519** (N=17) | **Other** (N=13) | *p* |
| **Mild** (N= 121) | 33(82.5%) | 0 | 0 | *na* | 56(35%) | 3(16.6%) | 7(18.4%) | 7(33.3%) | *ns* | 13(20%) | 1(12.5%) | 0 | 1(7.6%) | *ns* |
| **Severe** (N=113) | 2(5%) | 1(50%) | 1(50%) | ***0.03*** | 41(25.6%) | 8(44.4%) | 13(34.2%) | 10(47.6%) | ***0.05*** | 21(32.3%) | 3(37.5%) | 7(41.1%) | 6(46.1%) | *ns* |
| **Critical** (N=150) | 5(12.5%) | 1(50%) | 1(50%) | *ns* | 63(39.3%) | 7(38.8%) | 18(47.3%) | 4(19%) | *ns* | 31(47.6%) | 4(50%) | 10(58.8%) | 6(46.1%) | *ns* |
| *P* | ***<0.001*** | *ns* | *ns* |  | ***0.02*** | *ns* | ***0.02*** | *ns* |  | ***0.003*** | *ns* | ***0.003*** | *ns* |  |

*P* values were obtained from a Chi-square or Fisher exact test.

na: comparison not available

ns: non-significant

**Supplementary Figure S3. COVID-19 classification among patients infected with different lineages.** A. Barplot of patients infected with different lineages and it´s hospitalized status, controlled by age. B. Statistically significant comparisons between groups. P values were obtained from a Chi-Square Test.

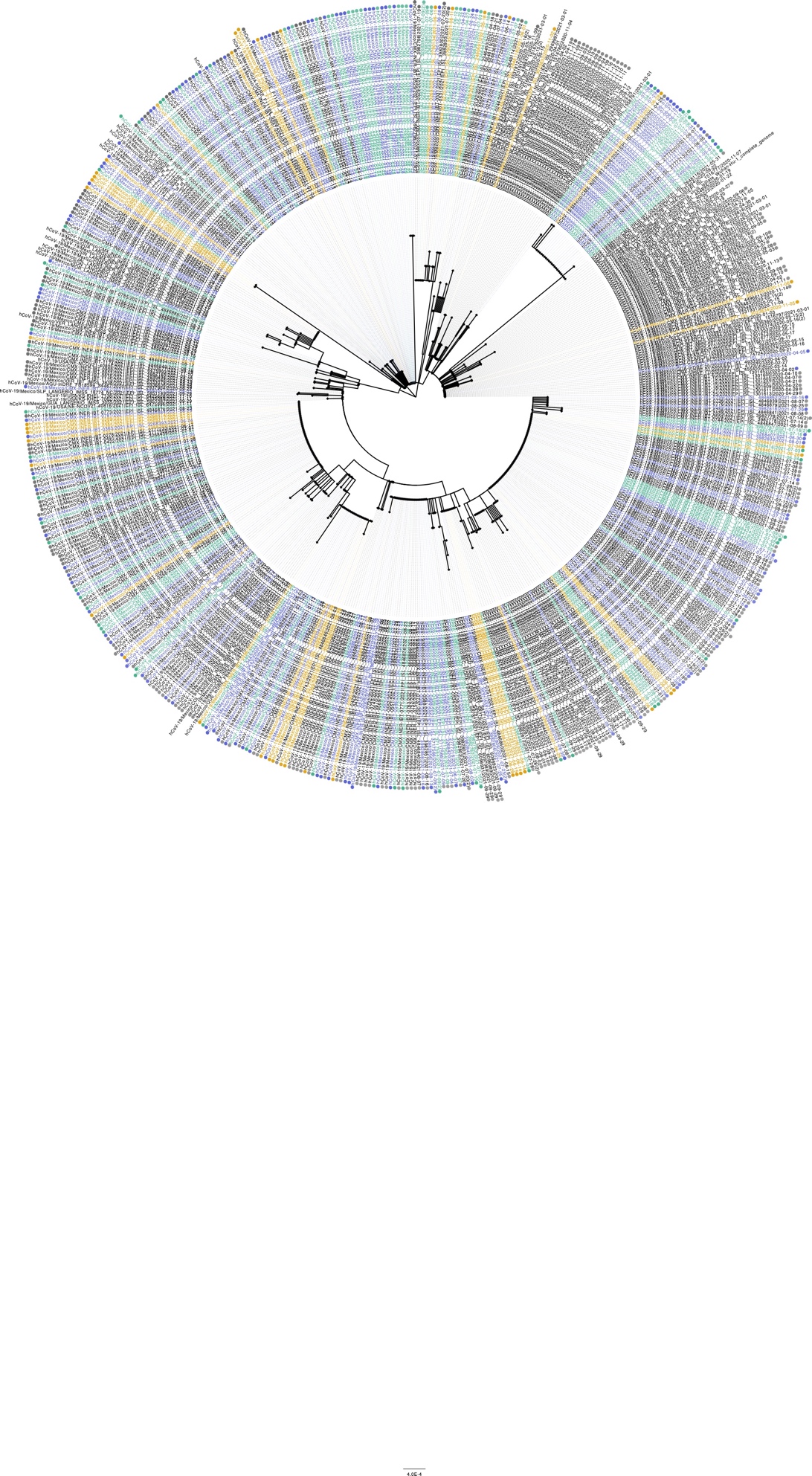

**Supplementary Figure S4. Phylogeny.** Maximum likelihood (ML) phylogenetic tree for the Spike whole sequence. ML tree from 541 sequences registered in GISAID was produced with 1,000 bootstrap replicates. The sequences obtained in this study are included. Branches are colored according to vaccination status; orange: unvaccinated, green: partially vaccinated, and purple: fully vaccinated patients.
